# Wastewater surveillance for COVID-19 response at multiple geographic scales: Aligning wastewater and clinical results at the census-block level and addressing pervasiveness of qPCR non-detects

**DOI:** 10.1101/2022.01.28.22269911

**Authors:** Hannah Safford, Rogelio E. Zuniga-Montanez, Minji Kim, Xiaoliu Wu, Lifeng Wei, James Sharpnack, Karen Shapiro, Heather Bischel

## Abstract

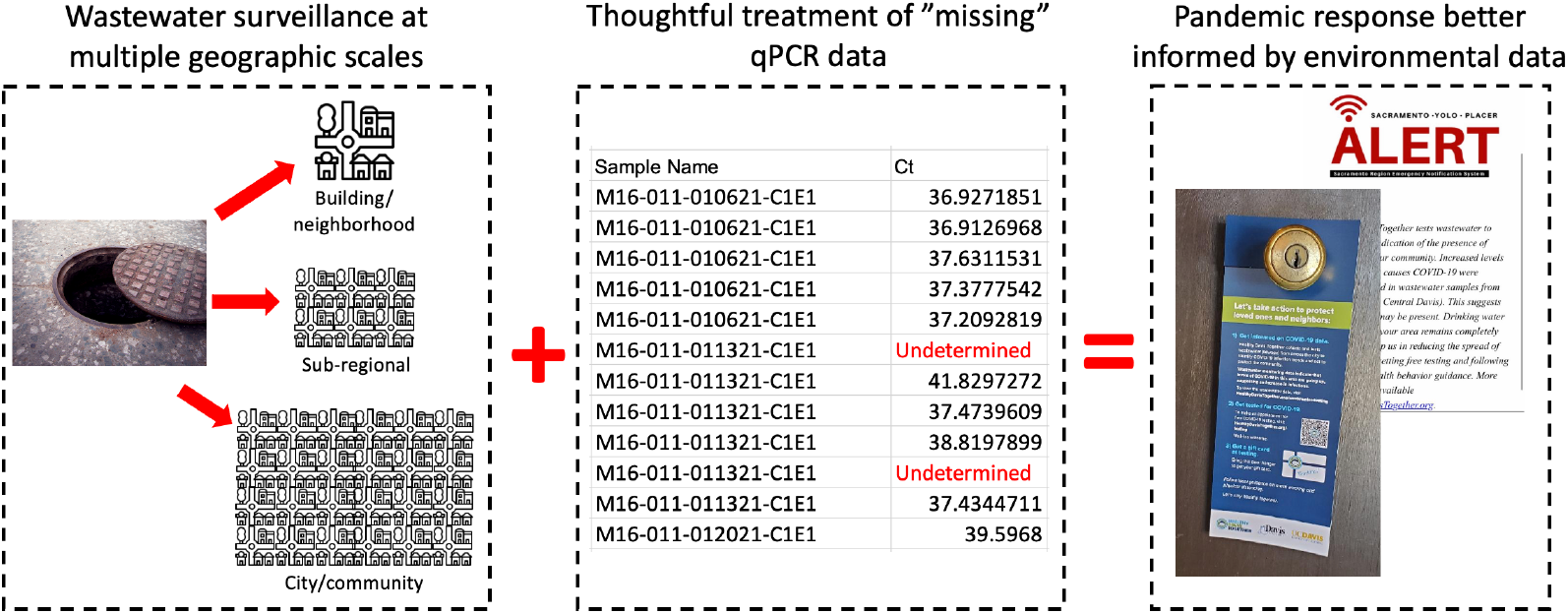

Wastewater surveillance is a useful complement to clinical testing for managing COVID-19. While good agreement has been found between community-scale wastewater and clinical data, little is known about sub-community relationships between the two data types. Moreover, effects of non-detects in qPCR wastewater data have been largely overlooked. We used data collected from September 2020–June 2021 in Davis, California (USA) to address these gaps. By applying a predictive probability model to spatially disaggregate clinical results, we compared wastewater and clinical data at the community scale, in 16 sampling zones isolating city sub-regions, and in seven zones isolating high-priority building complexes or neighborhoods. We found reasonable agreement between wastewater and clinical data at all scales. Greater activity (i.e., more frequent detections) in clinical data tended to be mirrored in wastewater data. Small, isolated clinical-data spikes were often matched as well. We also developed a method for handling such non-detects using multiple imputation and compared results to (i) single imputation using half the qPCR limit of detection, (ii) single imputation using maximum qPCR cycle number, and (iii) non-detect censoring. Apparent wastewater trends were significantly influenced by non-detect handling. Multiple imputation improved correlation relative to single imputation, though not necessarily relative to censoring.

## Introduction

Wastewater surveillance (also known as wastewater-based epidemiology, or WBE) has become widely recognized as a useful complement to clinical testing for managing COVID-19. Relative to large-scale diagnostic testing, wastewater surveillance offers a less resource-intensive way to monitor COVID-19 infections and spread among large numbers of people. Wastewater surveillance is also unbiased, capturing data on entire populations rather than just the subset of individuals who come in for clinical testing [1].

Most studies to date comparing wastewater and clinical data have focused on the community scale; i.e., comparing trends in data collected from the influent to a given wastewater treatment plant (WWTP) to trends in data collected from clinical tests of a subpopulation served by that WWTP. Such studies have frequently found good agreement between the two data sources. But little is known about relationships between wastewater and clinical data at sub-community levels. A first objective of this study was to advance and inform uses of wastewater surveillance at multiple scales for pandemic response. For instance, comparing data trends for wastewater collected from different neighborhoods could help public-health officials strategically allocate resources such as testing, contact tracing and vaccination outreach.

Separately, SARS-CoV-2 RNA in wastewater samples is typically quantified using either reverse transcription-quantitative polymerase chain reaction (RT-qPCR) or RT-droplet digital PCR (RT-ddPCR) [2]. While RT-ddPCR is becoming more popular for wastewater surveillance [3] due to its greater specificity and sensitivity [4,5], many laboratories continue to use RT-qPCR due to the higher cost and time requirements of RT-ddPCR and the large upfront capital investment of ddPCR instrumentation.

Bivins et al. (2021) recently drew attention to how variability in RT-qPCR methods and reporting affects results and interpretation [6]. An additional and important source of variability not considered by these authors is how non-detects are handled. qPCR non-detects occur routinely for reasons including low or zero starting target abundance, poor assay design/performance, or human error [7,8]. There is no current consensus on how to best manage qPCR non-detects. Researchers, whether through scientific software or manual analysis, typically handle non-detects either using <u>single imputation</u> (setting all non-detects equal to a constant value such as the mean of detected replicates, half the detection limit, or zero) or by <u>censoring</u> (excluding non-detects from analysis altogether) [8].

Unfortunately, both single imputation and censoring can substantially bias qPCR results [8]. The biasing effect is amplified when, as is often the case for wastewater data, the target is present in low concentrations to begin with. A second objective of this study was to demonstrate how different non-detect handling methods can affect apparent wastewater data trends, and to explore whether multiple imputation of non-detects can improve on more commonly used but less sophisticated approaches.

## Materials and Methods

### Study setting and design

We used wastewater data collected through the <u>Healthy Davis Together (HDT)</u> program in Davis—a small city of approximately 69,000 located in northern California—to (1) examine relationships between wastewater and clinical data at multiple spatial scales, and (2) explore the value of multiple imputation for handling qPCR non-detects.

HDT is a joint, multi-pronged initiative between the city of Davis and the University of California, Davis (UC Davis) for local management and mitigation of COVID-19. Beginning in November 2020, HDT made free, saliva-based PCR tests for COVID-19 available to anyone living or working in Davis. Uptake of the clinical-testing program was considerable. The fraction of Davis residents who reported receiving at least one COVID-19 test rose from 30% to 73% from September 2020 to March 2021. As of April 2021, Yolo County had performed the most tests per capita of California’s 58 counties, at a rate quadruple the state median.

HDT also conducts wastewater surveillance at the community, sub-regional, and building/neighborhood scales (Figure 1). At the <u>community scale</u>, samples are collected from the influent to the City of Davis Wastewater Treatment Plant (COD WWTP). The COD WWTP captures all of Davis’s municipal wastewater, with no contributions from UC Davis or from neighboring jurisdictions. At the <u>sub-regional scale</u>, samples are collected from sewershed nodes isolating the wastewater contributions of different geographic areas in the city. At the <u>building/neighborhood scale</u>, samples are collected from sewershed nodes isolating high-priority building complexes or neighborhoods identified through discussion with local officials. The HDT WBE program began in September 2020 with weekly samples collected from the COD WWTP. Zones were added and sampling frequency increased over the course of the sampling campaign (Figure S1). At full scale-up, the surveillance program sampled daily from the COD WWTP and 3x/week from each of 16 sub-regional and seven building/neighborhood zones.

**Figure 1.**
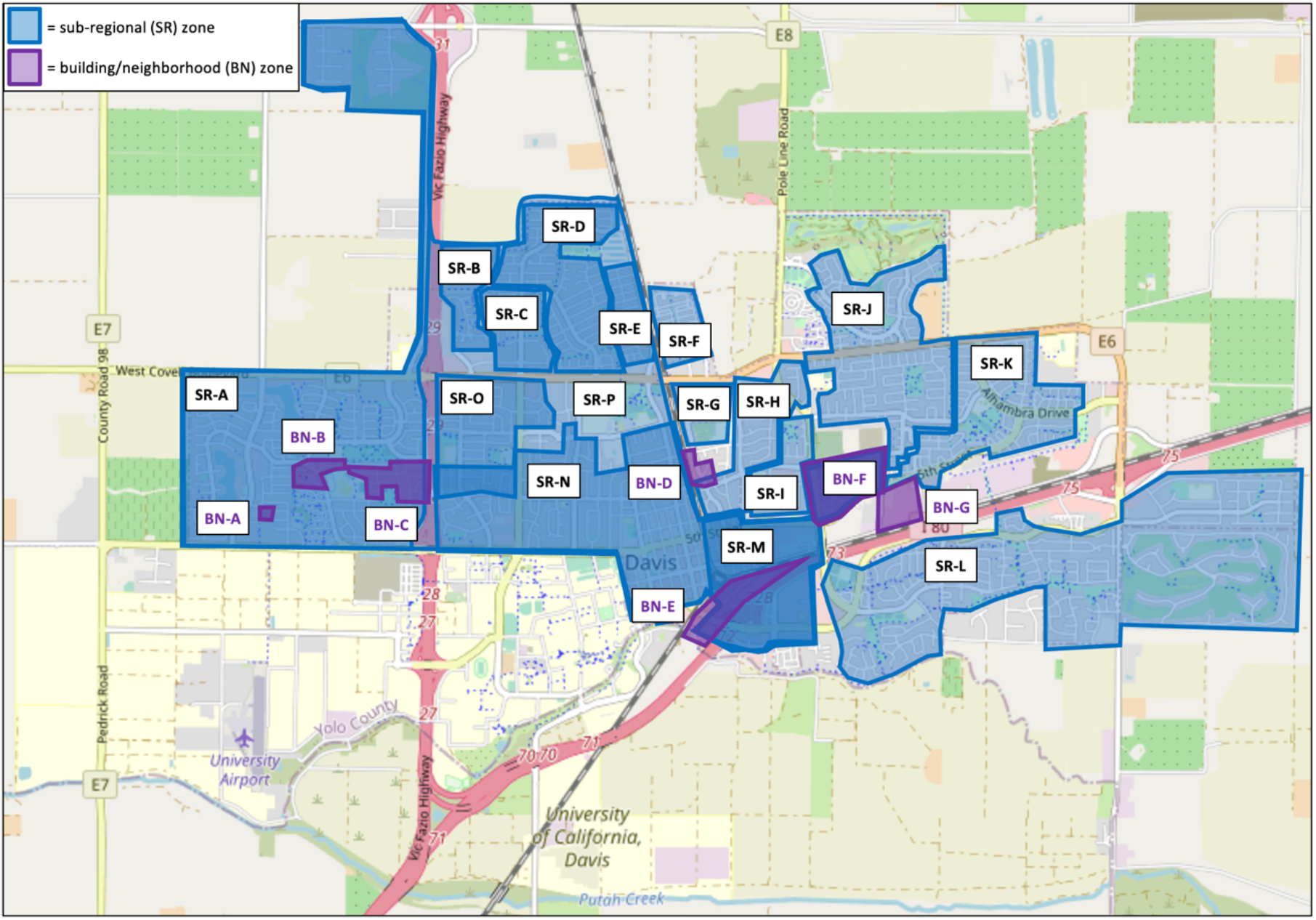
Map of sub-regional (SR; blue) and building/neighborhood (BN; purple) sampling zones for SARS-CoV-2 wastewater-based epidemiology in the city of Davis, CA. Note overlapping zones: in particular, zone SR-M overlaps the entirety of zone BN-F; zone SR-N overlaps a portion of zone SR-O and the entirety of zone SR-M; and zone SR-P overlaps the entirety of zones SR-A through SR-E as well as zones SR-O, SR-N, and SR-M.

### Sample collection

24-h composite samples were collected from each zone using insulated Hach™ AS950 Portable Compact Samplers (Thermo Fisher Scientific, USA) programmed to collect 30 mL of sample every 15 minutes. The bulk of samples were processed immediately, with a small number stored at 4°C for up to one week before processing.

### Sample processing

Samples were pasteurized for 30 minutes at 60°C to reduce biohazard risk while preserving RNA quality. Samples were then spiked with a known concentration of φ6 bacteriophage (strain HB104; generously provided by Samuel Díaz-Muñoz, UC Davis) as an internal recovery control [9,10]. The φ6 spike solution was prepared using previously described methods [11], modified slightly by using ATCC® Medium 129 in place of LB media. The final steps in the processing pipeline were sample concentration and extraction. From September 2020 through the end of February 2021, concentration was performed via ultrafiltration through 100 kDa Amicon® Ultra-15 centrifugal filter devices, and column-based extraction was performed manually using either the NucleoSpin^®^ RNA Stool Kit (Macherey-Nagel) or the AllPrep® PowerViral® DNA/RNA Kit (Qiagen). From February 2021 through June 2021, concentration was performed using Nanotrap® Magnetic Virus Particles (Ceres Nanosciences) and the MagMAX Microbiome Ultra Nucleic Acid Isolation Kit (Thermo Fisher) coupled with the KingFisher Flex liquid-handling system (Thermo Fisher). The particle-based method was far more conducive to automation and higher throughput than the ultrafiltration-based method, and the switch was necessary to accommodate greater numbers of samples as the sampling campaign scaled up.

We performed a four-sample comparison of the two methods and found that while the ultrafiltration method yielded higher concentrations of the fecal-strength indicator PMMoV, the magnetic-bead method appeared to be more sensitive for SARS-CoV-2, as indicated by detection of the N1 and N2 regions of the SARS-CoV-2 nucleocapsid gene (Figure S2; Table S1). Further details on the concentration and extraction protocols are available in *SI Materials and methods*. Raw data from the methods comparison is available in *SI Methods comparison*.

### RT-qPCR

Sample extracts were analyzed by one-step RT-qPCR for four targets: N1 and N2 targeting regions of the nucleocapsid (N) gene of SARS-CoV-2, φ6 bacteriophage, and pepper mild mottle virus (PMMoV; used for normalization of SARS-CoV-2 results). Per Bivins et al. (2021) [6], the Minimum Information for Publication of Quantitative Real-Time PCR Experiments (MIQE) checklist for this study is included as *SI MIQE*, and additional information on the RT-qPCR assay designs is available in Tables S1–S3. Triplicate wells were run for each target of each sample. Each run included a positive plasmid control and a no-template control, both run in duplicate. Six-point master standard curves for each target (Table S4) were constructed using serial dilutions of plasmid containing the targets at known concentrations, with each dilution assayed in triplicate or quadruplicate.

### Multiple imputation of non-detects

We developed and applied an expectation maximization-Markov chain Monte Carlo (EM-MCMC) model for multiple imputation of “missing” qPCR data: i.e., N1/N2 non-detects. Our multiple-imputation method for handling non-detects was inspired by the EM algorithm presented in McCall et al. (2014) [8]. Separately for each target (i.e., N1 and N2), we began by grouping results by sampling zone.^1^ Within each zone we modeled the Ct values (*X*_*i,t*_) for each technical replicate (index *i*) and sampling date (index *t*) as independent and identically distributed. The values are modeled with a normal distribution characterized by a common variance 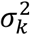 and common prior on the mean parameters *θ*_*i,t*_. The normal distribution is truncated such that it is positive.

We then used an empirical Bayesian approach to learn the prior for the model parameters. This approach enables discovery of hyperparameters shared by all samples from the same zone via the EM algorithm. The approach hence reduces variability in the inferred mean Ct values by specifying a common prior for all samples from a given location. Specifically, we modeled the priors for all *θ*_*i,t*_ and common *σ* as two Gamma distributions with shape and rate parameters 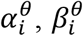 and 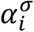, 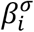 respectively. We estimated these hyperparameters^2^ with the EM algorithm, which alternates between calculating the posterior distribution for the latent (i.e., model-inferred) parameters given the current hyperparameters (E step) and updating the hyperparameters using maximum likelihood based on the posterior expectation. Because closed forms for the posterior distribution do not exist for this application, we sampled from the posterior using MCMC via Python’s Stan package (pystan).

The EM-MCMC algorithm can be summarized as:

(1) Initialize the hyperparameters 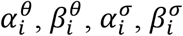.
(2) Generate *T* (a user-defined choice) Monte Carlo samples of the latent parameters *θ*_*i,t*_ and *σ* within the group using MCMC with the current hyperparameters.
(3) Compute the maximum likelihood estimates of the hyperparameters given the *T* sampled latent parameters (solved numerically via the scipy.stats.gamma.fit method).
(4) Repeat steps 2 and 3 until convergence of hyperparameters.

This process was carried out independently for each target and group. The Python script used for implementation is available at https://tinyurl.com/Safford-et-al-EM-MCMC.

### Data analysis

For multiple imputation, qPCR results grouped by zone were fed into the EM-MCMC model initialized with the hyperparameter priors 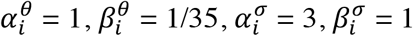. The model was run for 20 iterations, generating 10^4^ MCMC samples per iteration of which the first 500 were dropped. The model was then run again for one iteration (again with 10^4^ MCMC samples and 500 drop samples) using the hyperparameter estimates. The model output contained estimated posterior mean N1 and N2 Cts (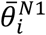 and 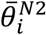) for each sample. the N1 and N2 targets for each location/date pair. Single imputation was performed for comparison method (i) by substituting 0.05 gene copies (gc)/reaction for N1 and 0.1 gc/reaction for N2 (i.e., half the N1 and N2 LODs calculated using 99% confidence level) as the target concentrations for any technical replicate yielding a non-detect. Single imputation was similarly performed for comparison method (ii) by substituting 0.010 gc/reaction and 0.047 gc/reaction (values calculated from the master standard curves using the assay’s maximum Ct of 45) as the target concentrations. Censoring was performed for comparison method (iii) by dropping non-detect values from N1 and N2 calculations.

N1, N2, and PMMoV reaction concentrations were converted to gc/L of initial sample based on effective volumes analyzed. MATLAB^®^ software (version R2021a; MathWorks) was used for subsequent analysis. N1 and N2 concentrations were averaged into a single concentration (*C*_*N*1*N*2_) per sample to facilitate data visualization and trend analysis. *C*_*N1N2*_ values were normalized using PMMoV according to the formula 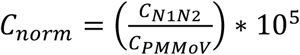, where 105 is a scaling factor. Normalized outliers were winsorized at the [1,95] percentile levels. Finally, relative normalized values were calculated separately for each non-detect handling method using the formula 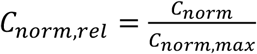, where *C*_*norm,max*_ is the maximum normalized value of all sewershed samples. Relative normalized values were used to visualize and compare trends in wastewater data processed using different non-detect handling methods. Because virus concentrations detected in WWTP influent differed substantially from virus concentrations detected in sewershed samples, these calculations were performed separately on sewershed and WWTP data. Values in between sampling dates were linearly interpolated to facilitate comparison of wastewater and clinical data, and the MATLAB “smoothdata” function was applied using a centered 7-day moving average.

### Probabilistic assignment of clinical data to sampling zones

All clinical data collected by HDT’s asymptomatic community-testing program^3^ since program inception were provided as an anonymized dataset indicating the date that each test was administered, the ZIP code and census block corresponding to the testee’s address, and whether the test was positive. Use of these data was deemed exempt from IRB review by the University of California, Davis IRB Administration. To compare clinical and wastewater data at the city/WWTP scale, we selected a subset of these data comprising all clinical-testing results for Davis ZIP codes (95616, 95617, and 95618). We designed a Python tool (available at https://tinyurl.com/Safford-et-al-Predictive) that combines information on municipal wastewater flows with U.S. Census Bureau data to probabilistically assign HDT asymptomatic testing results to sewershed sampling zones via three steps. First, we used the geospatial coordinates of all maintenance holes (MHs) in the Davis sewer system, along with information indicating the relative positions (upstream/downstream) of each MH, to build a graph capturing directional connections among all MHs (Figure 2A). Second, we used 2019 American Community Survey (ACS) data from the U.S. Census Bureau (UCSB) to estimate the number of people living in each census block included in the HDT clinical-testing dataset. We assume that each person in each census block produces the same amount of wastewater (a “unit”) each day, and that each person has an equal probability of discharging the wastewater unit to each MH located within the block (Figure 2B).

**Figure 2.**
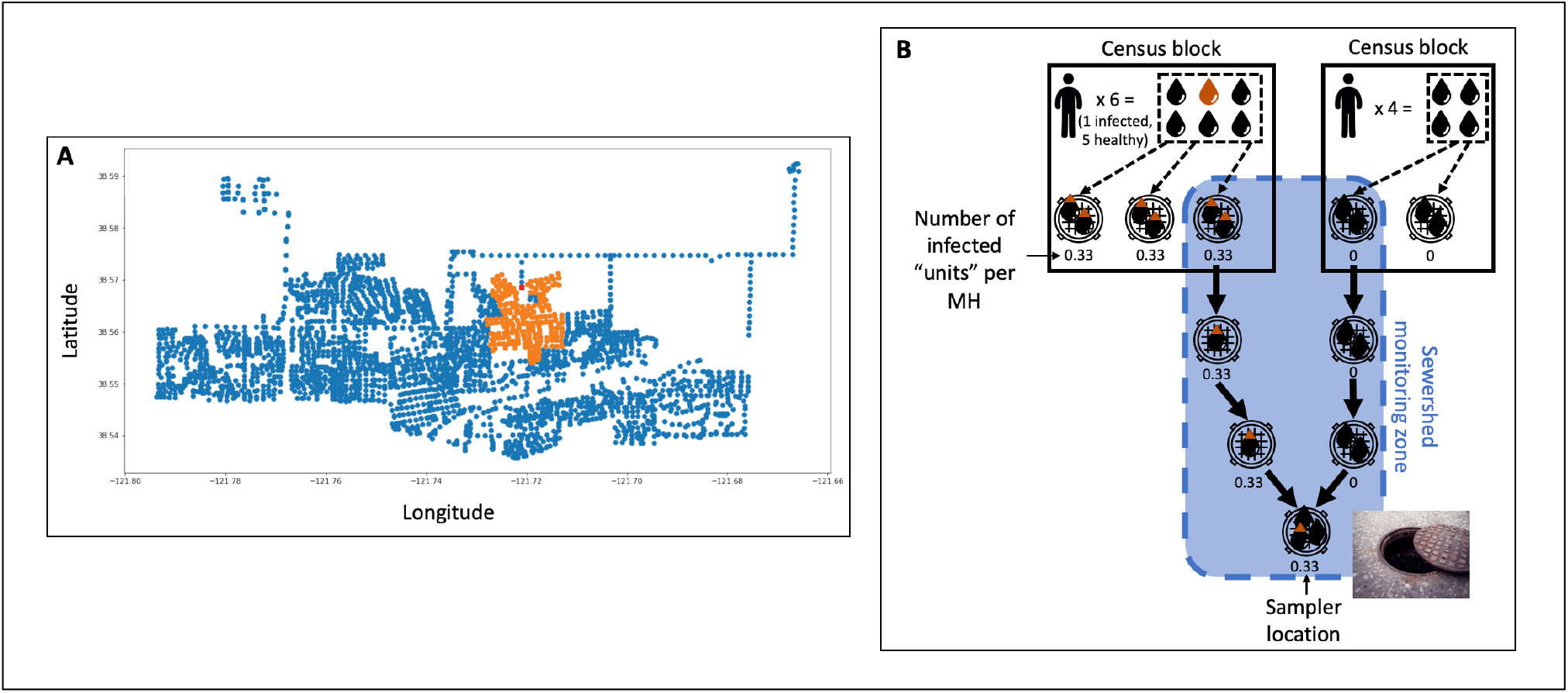
(A) Visualization of the connection graph showing all maintenance holes (MHs) in the City of Davis sewershed. Orange dots indicate all MHs upstream of a target MH (in red). (B) Illustration of how the connection graph is used to probabilistically assign positive clinical-test results from census blocks to sewershed monitoring zones for the purpose of comparing trends in wastewater data to trends in clinical data. In the illustration, the sewershed monitoring zone covered by the sampler location at bottom and indicated in blue spans two census blocks. The census block on the left has a population of six and one positive test result; the census block on the right has a population of four and no positive test results. Tracking flow through the connection graph results in a predicted 0.33 infections captured by the sampler.

Finally, we used the connection graph to probabilistically assign positive clinical-testing results from census blocks to sewershed monitoring zones. We excluded Zones SR-F, SR-G, BN-F, and BN-G from the probabilistic case assignment due to unreliable population data. Using the UCSB data, our tool estimated the populations of each of these zones to be less than 100: these estimates are unreasonably low given our *a priori* knowledge of the study setting. For Zones SR-F and BN-F, the unreliability could be because of the existence of potentially hard-to-count communities: several apartment complexes targeted at low-income renters in the former and a mobile-home park for senior citizens in the latter [12]. For Zones SR-F and BN-G, the unreliability can be attributed to the fact that the 2010 Decennial Census underlies the 2019 ACS. Zone SR-F comprises a large residential development that had not yet been built at the time that the 2010 Decennial Census was conducted. Zone BN-G largely comprises off-campus student housing that was under renovation at the same time. Population estimates for all other zones are provided in Table S7.

## Results and Discussion

### Sample collection and processing

We analyzed 964 wastewater samples collected during the sampling campaign, comprising 77 samples from the COD WWTP, 695 from the sub-regional zones, and 191 from the building/neighborhood zones. Overall, 204 samples (21%)—collected from September 24, 2020 through March 1, 2021—were processed using ultrafiltration + manual extraction and the remaining 760 (79%)—collected from February 24, 2021 through June 11, 2021—were processed using magnetic beads.

### Virus recovery and detection

Mean φ6 recovery was 1.30±0.28% across all samples, in line with values reported elsewhere [13]. At least one sample from each monitoring site and a total of 377 samples across all sites tested positive for SARS-CoV-2 (i.e., N1 or N2 above LOD in at least one technical replicate). Non-detect replicates were common even among positive samples; only 32 samples were positive for all N1 and N2 technical replicates.

N1 and N2 non-detect percentages were similar and inversely proportional to sampling scale (Table S5). This suggests that reliable detection of SARS-CoV-2 may become more challenging the further upstream in a sewershed that sampling is conducted. Pepper mild mottle virus (PMMoV) non-detects were never observed, indicating that the high percentages of N1/N2 non-detects can be attributed to frequently low abundance of SARS-CoV-2 in the wastewater samples rather than a systematic problem with the qPCR protocols used. This is further supported by (1) inclusion of N1 and N2 positive controls for every qPCR run, and (2) the fact that samples yielding higher numbers of positive technical replicates also exhibited lower Cts on average for those replicates (Table S6)—i.e., non-detects were more common when the target was present at lower concentrations.

### Multiple imputation of non-detects

Trace plots of posterior means generated by the EM-MCMC model over time generally showed good convergence. Trace plots of the MCMC samples exhibited no obvious patterns, indicating strong mixing of the Markov chains (Figure S3). Table 1 summarizes model output. The top half of the table shows that the number of positive replicates for a given sample exhibit a weak negative correlation with average standard deviations of N1 and N2 posterior mean Cts. This indicates that as the number of positive replicates increases, so too does the model’s confidence in its estimate of the “true” Ct. The bottom half of the table shows that the more positive replicates of a sample there are, the closer the average of those replicates is likely to be to the posterior mean Ct. The model output also shows that sampling scale exhibits a weak positive correlation with (1) standard deviations and (2) differences between the posterior mean and the average of positive replicates. This is likely because larger overall numbers of non-detects at higher-granularity sampling zones yield larger estimates for the per-zone variance. The very large posterior mean Cts for samples with zero positive replicates indicate that the model predicts that these samples contain essentially zero SARS-CoV-2 RNA.

**Table 1.** Summary of imputation model output

| Average imputed standard deviation |  |  |  |  |  |  |  |  |
| --- | --- | --- | --- | --- | --- | --- | --- | --- |
| Sampling scale | N1* |  |  |  | N2* |  |  |  |
|  | Number of positive replicates |  |  |  | Number of positive replicates |  |  |  |
|  | 0 | 1 | 2 | 3 | 0 | 1 | 2 | 3 |
| Community | 7.85<br>(0.10) | 3.27<br>(0.05) | 2.85<br>(0.07) | 2.71<br>(0.03) | 7.84<br>(0.09) | 3.29<br>(0.14) | 2.91<br>(0.09) | 2.73<br>(0.04) |
| Sub-regional | 11.22<br>(3.89) | 3.90<br>(0.81) | 3.28<br>(0.57) | 2.92<br>(0.36) | 11.30<br>(3.85) | 3.79<br>(0.66) | 3.27<br>(0.53) | 2.83<br>(0.33) |
| Building/neighborhood | 20.15<br>(4.15) | 4.69<br>(0.90) | 3.42<br>(0.57) | 2.94<br>(0.28) | 20.01<br>(4.48) | 4.25<br>(0.64) | 3.69<br>(0.62) | 3.20<br>(0.50) |
| Overall | 13.78<br>(5.83) | 3.99<br>(0.87) | 3.24<br>(0.55) | 2.89<br>(0.33) | 13.73<br>(5.78) | 3.79<br>(0.66) | 3.25<br>(0.53) | 2.86<br>(0.35) |
| Average difference in mean Cts |  |  |  |  |  |  |  |  |
| Sampling scale | N1* |  |  |  | N2* |  |  |  |
|  | Number of positive replicates |  |  |  | Number of positive replicates |  |  |  |
|  | 0 | 1 | 2 | 3 | 0 | 1 | 2 | 3 |
| Community | 13.66<br>(0.06) | 8.29<br>(0.51) | 3.82<br>(1.29) | 1.53<br>(0.17) | 13.64<br>(0.06) | 6.89<br>(1.20) | 3.86<br>(0.55) | 1.41<br>(0.15) |
| Sub-regional | 20.47<br>(7.38) | 8.99<br>(1.49) | 4.73<br>(1.26) | 1.96<br>(1.02) | 20.62<br>(7.31) | 7.97<br>(1.34) | 4.37<br>(0.87) | 1.58<br>(0.58) |
| Building/neighborhood | 37.63<br>(7.94) | 10.26<br>(1.45) | 4.94<br>(0.87) | 1.88<br>(0.96) | 37.37<br>(8.48) | 8.41<br>(1.23) | 4.98<br>(0.78) | 1.76<br>(0.63) |
| Overall | 25.37<br>(11.18) | 9.16<br>(1.52) | 4.65<br>(1.26) | 1.89<br>(0.93) | 25.27<br>(11.07) | 7.90<br>(1.36) | 4.34<br>(0.85) | 1.57<br>(0.55) |

\*Upper value indicates average; lower (parentetical) value indicates standard deviation.

### Comparison of clinical and wastewater data

Davis is a small community that experienced a relatively low COVID-19 burden during this study, daily numbers of HDT-reported cases were generally low. Double-digit numbers of confirmed cases were reported on only 11 of the 234 days included in this study, and days on which the number of confirmed cases was zero or one were common. Probabilistically assigned case levels at the sub-regional and building/neighborhood scales were frequently fractional and near zero as a result. Figure S4 co-plots the clinical data and normalized SARS-CoV-2 concentrations for each sampling zone.

Correlations between clinical and wastewater data were reasonably good at all sampling scales. Zones and time periods exhibiting greater activity (i.e., more frequent detections) in clinical data tended to also exhibit greater activity in wastewater data. We generally observed more data activity at the community and sub-regional scales than at the building/neighborhood scale, as well as more activity in bigger zones at a given scale. This finding is logical—average COVID-19 case counts will be higher in zones covering more people—but important because it indicates that the predictive probability model is reasonably successful at assigning positive cases to the appropriate sampling zones.

In multiple zones (e.g., BN-D, BN-E, SR-C, SR-E, and SR-I), even relatively small and isolated spikes in clinical data were matched by spikes in wastewater data. As Zulli et al. (2021) observe, parallel spikes in wastewater virus concentrations and clinical case rates recorded at the community and regional levels during the winter 2020/2021 COVID-19 surge indicate that wastewater monitoring can provide accurate information on changes in disease burden [14]. Our results provide evidence that wastewater monitoring is similarly valuable at the sub-regional and building/neighborhood levels.

Separately, wastewater data from most zones sampled in this study were characterized by major peaks and valleys—with a high positive result frequently occurring right after a low positive result and vice versa—rather than smooth trends. This phenomenon can be mostly attributed to low-frequency sampling during the period of highest disease burden. Based on daily sampling of wastewater from multiple WWTPs in Wisconsin, Feng et al. (2021) concluded that “a minimum of two samples collected per week [is] needed to maintain accuracy in trend analysis” [15]. Due to staffing and lab-capacity constraints, however, wastewater samples for this study were only collected on a weekly basis from November through late January. Trend smoothness generally improved when sampling frequency was increased in late winter / early spring. Data from zone SR-L provide a particularly good example of how increased sampling frequency made it easier to trace trends.

Even after sampling frequency increased, we occasionally observed isolated high-positive results that did not appear part of broader trends (e.g., for zone SR-H in late March and zone SR-F in late April). These isolated positives could be due to aberrations (such as an infected group of individuals temporarily visiting a zone or coincidental passage of a large amount of virus-rich fecal matter near an autosampler actively drawing up volume) rather than sustained community spread. This possibility cautions against basing public-health interventions on individual data points.

Occasional mismatches between wastewater and clinical data trends (e.g., the spike observed for clinical—but not wastewater—data in early April for Zone SR-B) have multiple possible explanations. One is that while the predictive probability model performs reasonably well, it is still at best an approximation of the number of clinically confirmed cases in each wastewater sampling zone. Furthermore, generally low COVID-19 levels in Davis yielded sparse and/or weak positive signals in the clinical data, which in turn made it difficult to perceive trends at more granular spatial levels. A more precise comparison of wastewater and clinical data would require disclosing the addresses of individuals testing positive—an unacceptable privacy violation.

A second explanation is that the HDT dataset used in this study is incomplete. The dataset does not include results from other COVID-19 testing opportunities available to Davis residents (e.g., tests conducted in medical settings or through county-run testing programs). The HDT dataset also does not include results from the parallel on-campus testing program for UC Davis students and employees even though these individuals frequently reside off campus. This explanation could account for the February spike in wastewater—but not clinical—data observed for Zone BN-D, since Zone BN-D includes an apartment complex targeted at students.

A final explanation is that neither WBE nor clinical testing reliably capture the “true” level of COVID-19 infections in a sampling zone. WBE results can be affected by many factors, including variability in SARS-CoV-2 excretion rates [16], wastewater composition and temperature, average in-sewer travel time, per-capita water use [17], autosampler settings [18], and movement of people in and out of sampling zones. Clinical-testing results can be further biased by various types of self-selection [19,20]. Though it is impossible to precisely determine the relative contributions of these factors and biases, context can suggest which are likely to have the greatest influence in a given instance. For example, an unexplained spike in wastewater—but not clinical—data for a zone housing disproportionate numbers of individuals with characteristics that could cause lower propensity to test (e.g., limited access to transportation; low English proficiency) could be a sign of the presence of infected individuals detected through WBE but not clinical testing.

### Comparison of non-detect handling methods

We compared our multiple-imputation method with three other, commonly used methods for handling non-detects in wastewater qPCR data:

1. **[LOD**_**0.5**_]. Single imputation with half the detection limit.
2. **[Ct**_**max**_]. Single imputation with the maximum qPCR cycle number.
3. **[Ct**_**avg**_]. Censoring non-detects entirely (and setting average concentrations of samples with no positive replicates to zero).

Figure S5 co-plots the clinical data and relative normalized SARS-CoV-2 concentrations calculated using each method for each sampling zone; Figure 3 provides a representative subset of these plots.

**Figure 3.**
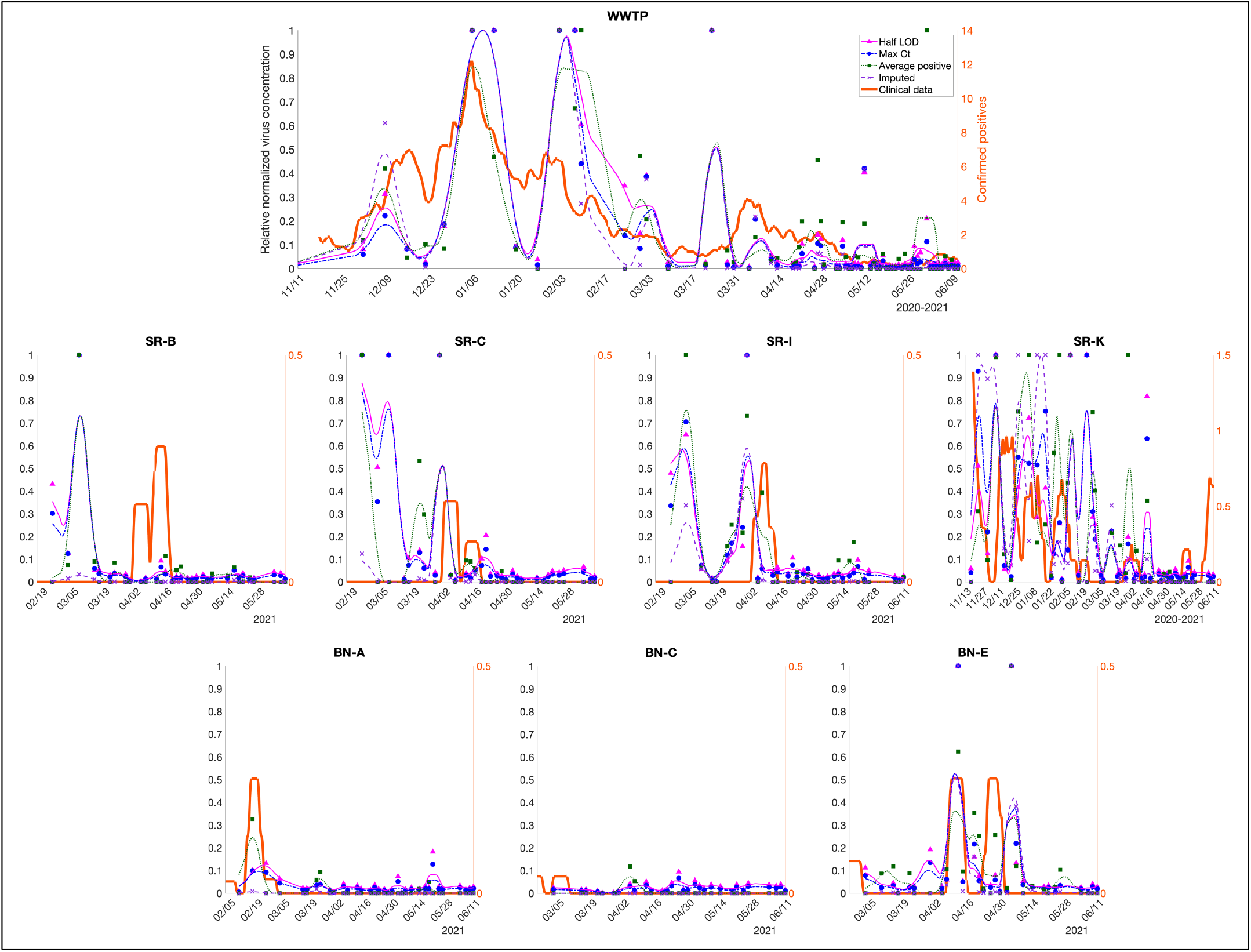
Wastewater vs. clinical data in Davis, showing effects of different methods of handling non-detects. Symbols represent individual sample results; lines represent trends (as centered 7-day moving averages). Representative suite of plots; see Figure S6 for plots from all zones.

The plots show that trends in normalized virus concentration can be substantially influenced by the way in which non-detects are handled. For instance, peak relative normalized virus concentrations in the WWTP data are much higher when calculated using multiple imputation than when calculated using any of the comparison methods. Conversely, relative normalized virus concentrations in samples collected at zones SR-B, SR-C, and SR-D from mid-February through mid-March are much lower when calculated using multiple imputation. These zones are all geographically proximate and of roughly equivalent size, indicating that zones with similar characteristics may be similarly susceptible to bias in non-detect handling method.

We applied Spearman’s rank-order correlation to quantitatively assess how well trends captured by multiple imputation and the three comparison methods match clinical-data trends. Results are summarized in Table 2. While the Spearman correlation analysis provides a framework for interpreting the data, it suffers from several limitations. These include factors discussed above, including the low COVID-19 burden in Davis, imprecision associated with the predictive probability model, and external factors that can inherently confound wastewater analysis. It is also hard to definitively “match” trends in clinical and wastewater data. For instance, trends in clinical data collected from symptomatic individuals been observed to lag trends in wastewater data [21,22]. But it is unknown whether and to what extent this lag may apply when clinical data derives from a large-scale asymptomatic testing program like HDT.

**Table 2.**
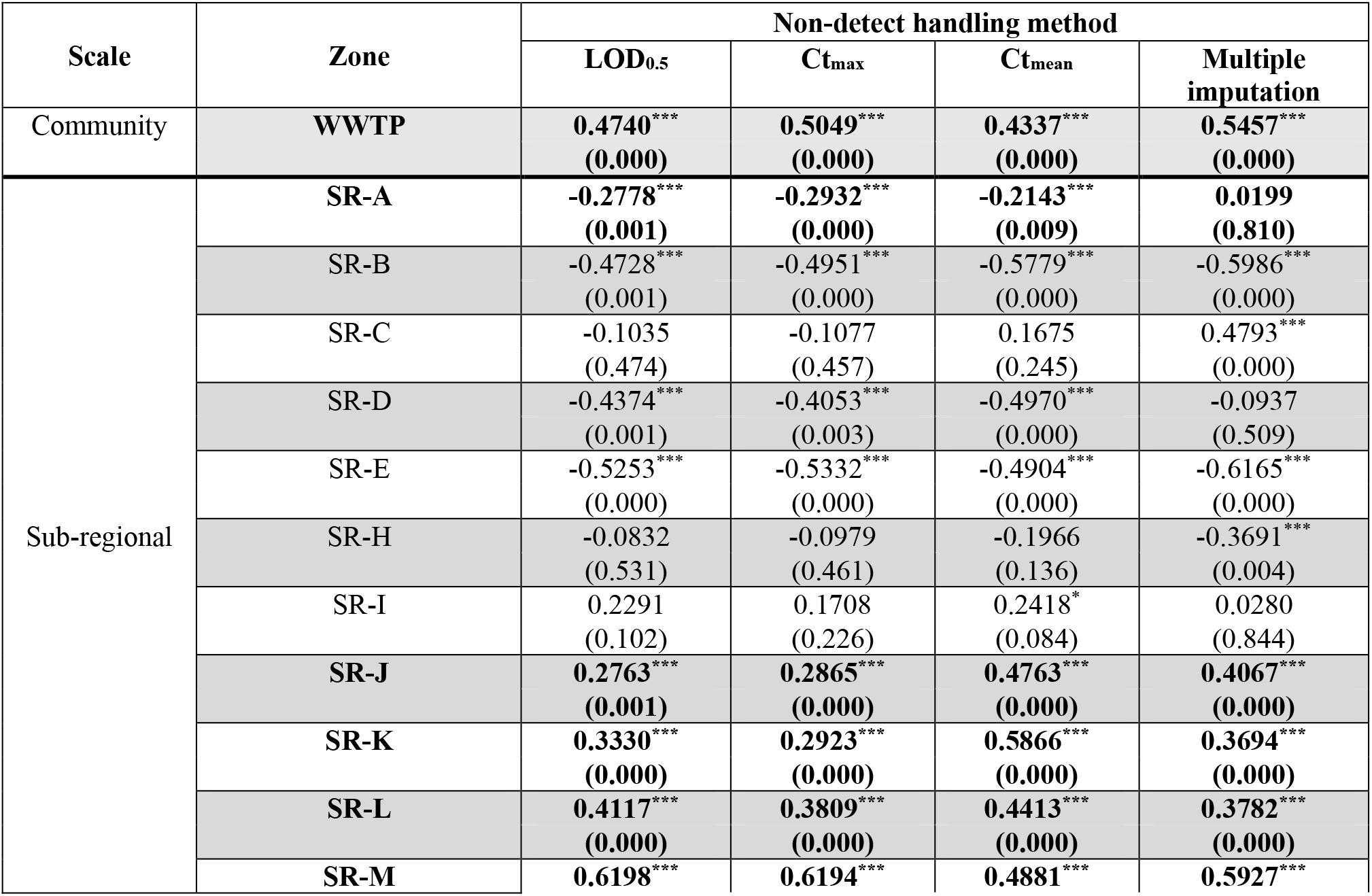
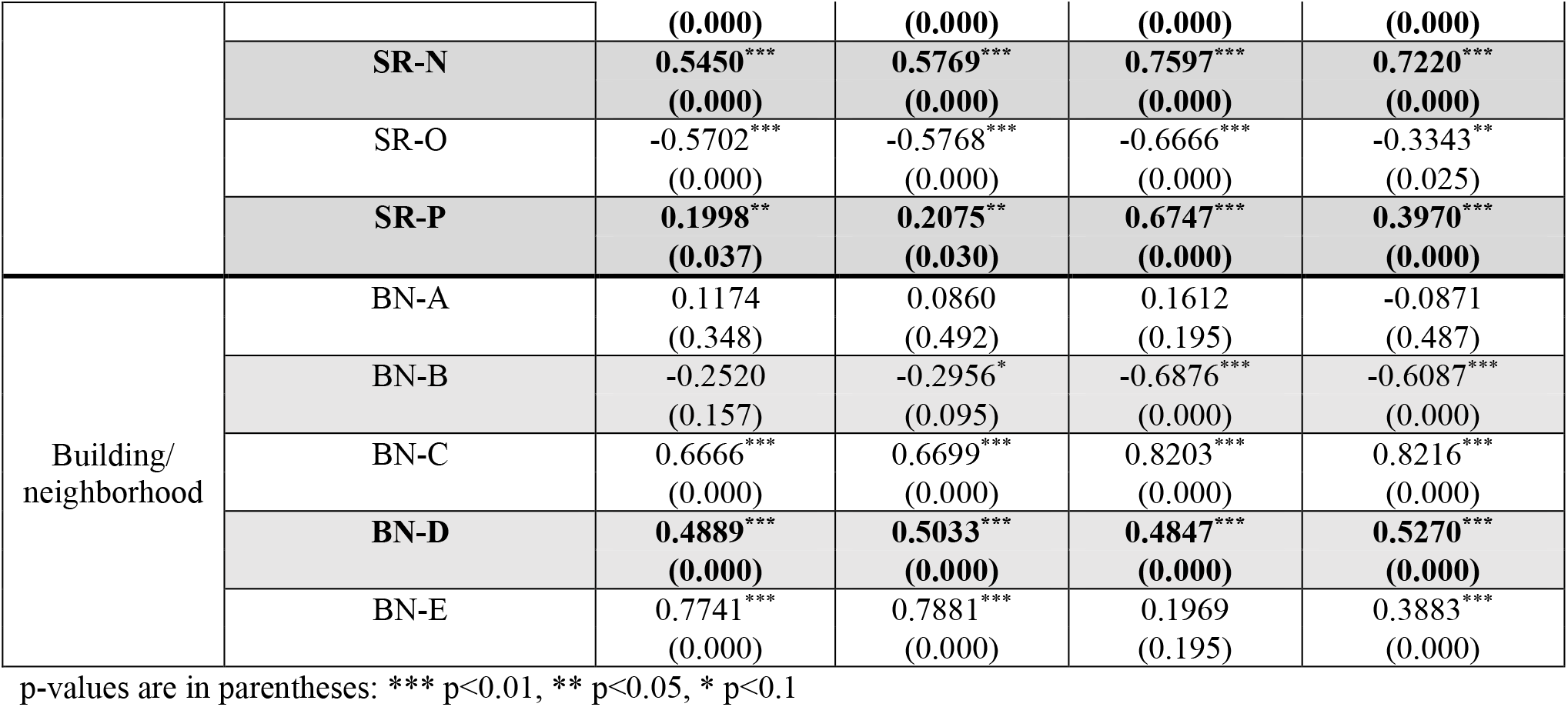
Spearman’s rank-order correlation coefficients between clinical cases and normalized virus concentration, by zone and non-detect handling method. Bolded rows indicate zones where wastewater surveillance began prior to the winter COVID-19 surge. As described in the text, Zones SR-F, SR-G, BN-F, and BN-G were excluded from the clinical/wastewater data comparison.

With these caveats in mind, important takeaways from the correlation analysis are as follows. First, because the LOD_0.5_ and Ct_max_ methods involve a similar approach, their correlation coefficients track more closely with each other than with the Ct_avg_ or multiple imputation coefficients.

Second, much higher correlation coefficients were generally observed for the 11 zones where wastewater surveillance began prior to the winter COVID-19 surge. This can be explained by greater activity in the wastewater and clinical data during the winter surge, as well as by the fact that sampling zones added later in the campaign were generally smaller—and hence less active— than zones added earlier. Time periods and zones with more data activity provide more positive data points on which to perform meaningful rank-order comparisons. The larger datasets available for zones where sampling began early also strengthen the robustness of data comparisons (as indicated by the universally low p-values of correlation coefficients for these zones). Correlation coefficients calculated for time periods and zones with less activity and where sampling began later can be easily skewed by a small number of disparate results between the wastewater and clinical data.

Third, the correlation analysis revealed no clear “winner” among the four non-detect handling methods. For the 11 zones where sampling began early, correlation coefficients tended to be weaker for the LOD_0.5_ and Ct_max_ methods. The Ct_avg_ method performed the best on average across these zones, but also performed the worst (with multiple imputation performing best) for the WWTP data—the one zone where imprecision associated with probabilistic assignment of confirmed positives was not a factor, and hence where correlation between the wastewater and clinical data is arguably most meaningful.

### Implications for multiscale deployment of wastewater surveillance

This study demonstrates that wastewater surveillance can provide a useful complement to clinical testing at multiple levels of spatial granularity. Visual and quantitative comparison of data from HDT’s universal asymptomatic clinical-testing program with data from the initiative’s intensive wastewater-surveillance campaign revealed meaningful correlations at the community, sub-regional, and building/neighborhood scales. The predictive probability model we developed for disaggregating deidentified HDT case data by wastewater-sampling zone provides a framework that can be easily extended to support sub-community comparison of clinical and wastewater data in other settings.

This study also demonstrates that qPCR non-detects are an important but often overlooked determinant of apparent trends in wastewater data. Our results indicate that single imputation with a value equivalent to half the assay limit of detection, the maximum qPCR cycle number, or similar generally weakens correlation between wastewater and clinical data. Simply censoring non-detects seemed to be a better approach. We did not find clear evidence that multiple imputation of non-detects delivers consistently better correlations between wastewater and clinical data than censoring does. However, the fact that multiple imputation yielded the strongest correlation for the WWTP data suggests, as discussed above, that this approach merits further investigation. With adjustments to the algorithm, tuning parameters, and variable groupings used herein, multiple imputation could support more reliable analysis of trends in wastewater data—and hence position wastewater surveillance as an even stronger weapon in the fight against pandemic spread.

We acknowledge two limitations of our work. First, some comparisons presented herein are incomplete because sampling zones were added over time. Only two of the seven sampling zones at the building/neighborhood scale, for instance, were active during the winter pandemic surge. Though this means that our results do not provide deep insight into the value of spatially granular wastewater surveillance during periods of peak disease spread, we note that wastewater surveillance tends to be more valuable outside of such periods—e.g., as an early-warning system when background case levels are low. Second, we did not rigorously test the effect of different data groupings when running the multiple-imputation model. Though grouping data by sampling zone is a logical choice, it is possible that alternate groupings (e.g., grouping by sampling scale, grouping temporally, pooling results from adjacent sites, etc.), coupled with appropriate tuning of model parameters, could significantly alter and perhaps improve results. Indeed, further refinement and optimization of our multiple-imputation model is needed to unlock the full potential of this promising approach to handling qPCR non-detects in wastewater data.

## Supporting information

SI MIQE

SI Sample data and metadata

SI Methods comparison

## Data Availability

All data produced in the study are either provided in the SI or available upon reasonable request to the authors.

## Supporting Information

The following files are available free of charge.

- Additional materials and methods, including information on qPCR assay, and methods comparison, and sampling zone populations (SI Materials and methods, figures, tables.pdf)
- Raw data and metadata from sample collection and analysis (SI Sample data and metadata.xlsx)
- Probabilistic assignments of clinical-testing results (SI Clinical model output.xlsx)
- Raw data from methods comparison (SI Methods comparison.xlsx)
- MIQE checklist (SI MIQE.xls)

## Present Addresses

N/A

## Author Contributions

H. Bischel and K. Shapiro oversaw the study; M. Kim tested and optimized qPCR assays; H. Safford and R. Zuniga-Montanez performed lab work; X. Liu and J. Sharpnack designed the multiple imputation model; L. Wei and J. Sharpnack designed the predictive probability model; H. Safford analyzed data, generated figures, and drafted the manuscript; R. Zuniga-Montanez, M. Kim, J. Sharpnack, K. Shapiro, and H. Bischel contributed text and edits to the final manuscript. All authors have given approval to the final version of the manuscript.

## Notes

N/A

## Acknowledgements

We acknowledge the invaluable contributions of the following individuals: L. Rueda, W. Bess, R. Pechacek, and M. Clauzel for lab support; N. Krasner for data cleanup and entry; S. Gryzcko, A. Livingston, S. Macomb, and J. Miller for coordinating sample collection; M. Nuno for input on modeling and statistical analysis. This project was carried out with the generous support of Healthy Davis Together.

## Briefs

N/A

## Synopsis

Analysis of how multiscale wastewater surveillance can inform pandemic response and protect public health.

## Footnotes

1 The method can accommodate other types of groupings—e.g., by sampling scale.

2 A hyperparameter is a parameter used only to influence the learning behavior of a model. Hyperparameter values are not derived from training or experimental data. By contrast, parameters are values determined by the model from analyzing input data.

3 UC Davis also conducts a testing program open only to UC Davis students and employees. Data from this program were not included in the dataset used for this study.

