## Supplementary material for "Wastewater surveillance for COVID-19 response at multiple geographic scales: Aligning wastewater and clinical results at the census-block level and addressing pervasiveness of qPCR non-detects": SI Materials and methods, figures, tables

### Supplementary Materials and Methods

*Processing method 1: Ultrafiltration + column-based extraction*

In the ultrafiltration method, 50-mL aliquots of pasteurized wastewater sample were spiked with φ6 bacteriophage solution containing $3.51 \times 10^8$ gene copies (gc) per uL of solution. Early in the sampling campaign the spike volume was 18 µL; this volume was decreased to 5 µL later on. Recovery calculations accounted for differences in spike volume, and spike volumes for each sample are recorded in *SI Data*. Spiked aliquots were vigorously shaken by hand and then incubated for 30 minutes at room temperature. Following incubation, the ultrafiltration method followed a protocol based on Ahmed et al. (2020).[1] Sample aliquots were centrifuged for 10 minutes at 4°C and 4,000 rcf to settle out large solids. 100 kDa Amicon® Ultra-15 centrifugal filter devices (Fisher Scientific) were pre-wet with 10 mL of autoclaved 1x Tris-EDTA (TE) buffer. 15 mL of sample supernatant was loaded into each device, and devices were centrifuged at 4°C and 4,000 rpm for as long as it took to pass the entire volume of sample through the device. Flow-through was discarded, and devices were reloaded with additional sample twice more to concentrate the entire 45 mL of supernatant volume. In the event of filter clogging for a challenging sample, the entire contents of the upper reservoir of the Amicon® device were transferred into a new pre-wetted device and concentration was resumed. The retentate was then augmented with autoclaved 1x TE buffer to achieve a known final volume of 1 mL. A small subset of samples was augmented to only 0.5 mL, and an additional subset was augmented to a volume ranging from 1–2 mL. Disparate volumes were accounted for during data analysis.

Samples concentrated through ultrafiltration were extracted using the NucleoSpin® RNA Stool Kit (Macherey-Nagel). Due to supply-chain limitations, the AllPrep® PowerViral® DNA/RNA Kit (Qiagen) was substituted for a small number of extractions (noted in *SI Data*). The NucleoSpin® and AllPrep® PowerViral® kits involve similar approaches and internal tests yielded comparable results. A 200 µL subsample from the ultrafiltration concentrate was always used as the starting sample volume. Macherey-Nagel kit was used following the manufacturer's instructions for isolating total RNA with the following modifications: (1) bead beating was only carried out for 2 minutes, and (2) the DNA digestion step was omitted. The Qiagen kit was used following the manufacturer's instructions but omitting the bead-beating step. With both kits, samples were eluted into 105 µL of RNAse-free water. Extracts were typically stored on ice and subjected to RT-qPCR analysis the same day to avoid losses from RNA degradation. When same-day analysis was not possible, extracts were immediately stored at -80°C until analysis (noted in *SI Data*).

*Processing method 2: Particle-based capture*

In the particle-based capture method, 5 mL of each wastewater sample was deposited into a separate well of a KingFisher 24 deep-well plate (Thermo Fisher). Each well was spiked with 5 µL of φ6 bacteriophage solution containing $9.02 \times 10^7$ to $3.51 \times 10^8$ gc per uL of solution. Each spiked sample was manually agitated by pipetting up and down using a 5-mL pipette at least three times; samples were then incubated for 30 minutes at room temperature. Following incubation, concentration was carried out using Nanotrap® Magnetic Virus Particles (Ceres Nanosciences) on a KingFisher Flex robot (Thermo Fisher). Concentration followed the protocol by Karthikeyan et al. (2021),[2] but with only 5 mL instead of 10 mL starting sample volume. Concentrated viruses were eluted from the Nanotrap® beads using 400 mL of lysis buffer per sample from the MagMAX Microbiome Ultra Nucleic Acid Isolation Kit (Thermo Fisher). Concentrated samples were extracted using the MagMAX kit in conjunction with 96 deep-well

[1] Ahmed, W.; et al.. (2020). Comparison of virus concentration methods for the RT-qPCR-based recovery of murine hepatitis virus, a surrogate for SARS-CoV-2 from untreated wastewater. *Science of The Total Environment*, 739.
[2] Karthikeyan, S.; et al. (2021). High-Throughput Wastewater SARS-CoV-2 Detection Enables Forecasting of Community Infection Dynamics in San Diego County. *mSystems*, 6(2).

plates on the KingFisher Flex, per the manufacturer's recommendations. Samples were eluted in 100 μL of elution solution from the MagMAX kit. Again, extracts were typically stored on ice and immediately subjected to same-day analysis. When same-day analysis was not possible, extracts were immediately stored at -80°C until analysis (noted in *SI Data*).

*RT-qPCR*

RT-qPCR amplifications were performed in 25 μL reactions on StepOnePlus qPCR thermocyclers (Applied Biosystems). Each reaction contained the following components: 0.625 μL bovine serum albumin (BSA; 25 mg/mL), 1.875 μL primer/probe mix, 2.5 μL RNAse-free water, 2.5 μL 10x Multiplex Enzyme Mix from the Path-ID™ Multiplex One-Step Kit (Applied Biosystems), 12.5 μL of 2x Multiplex RT-PCR Buffer from the Path-ID™ kit, and 5 μL of sample extract or control. Preparation and plating of RT-qPCR mastermix was carried out in a separate location from sample loading to avoid contamination.

Table S1 summarizes primers, probes, and cycling conditions for RT-qPCR assays performed as part of this work; Table S2 provides the primer/probe mix recipes; and Table S3 details the master standard curves used for each target.

*Method comparison*

The methods comparison was performed on four raw samples: three from different sewershed sampling sites, and one from the City of Davis WWTP. Samples were pasteurized on arrival, stored at -80°C for several weeks, and then thawed at room temperature prior to processing. Each of the four samples was processed using one of two methods: ultrafiltration or magnetic beads. The ultrafiltration method was performed as described above, using the NucleoSpin® kit for column-based extraction. Because the methods comparison was performed prior to our lab's acquisition of a KingFisher Flex, the magnetic-bead method was carried out manually, according to a protocol adopted from Rasile and Maas (2021).[3] In brief, 600 μL of Nanotrap® particles were added to 40 mL of each sample in 50-mL conical tubes. Samples were inverted several times and incubated for 20 minutes at room temperature. Sample tubes were placed on magnetic racks for 20 minutes to collect the particles, and the supernatant was discarded. Particles were resuspended in 1 mL of lysis buffer from the PureLink™ Viral RNA/DNA Mini Kit (Invitrogen), and the entirety of the suspension was then extracted using the PureLink™ kit according to the manufacturer's instructions, but without the addition of carrier RNA. RT-qPCR was then performed as described above. The method comparison employed three process replicates per method per sample, and three RT-qPCR technical replicates per process replicate. Raw data from the method comparison is available in *SI Method comparison*, and results are summarized in Figure S2 and Table S1.

[3] B. Rasile, K. Maas, SARS-CoV-2 Wastewater RNA Concentration and Extraction (Nanotrap and NucleoMag® RNA Water). Protocols.io. Available at dx.doi.org/10.17504/protocols.io.bn58mg9w. Deposited 19 January 2021.

### Supplementary figures

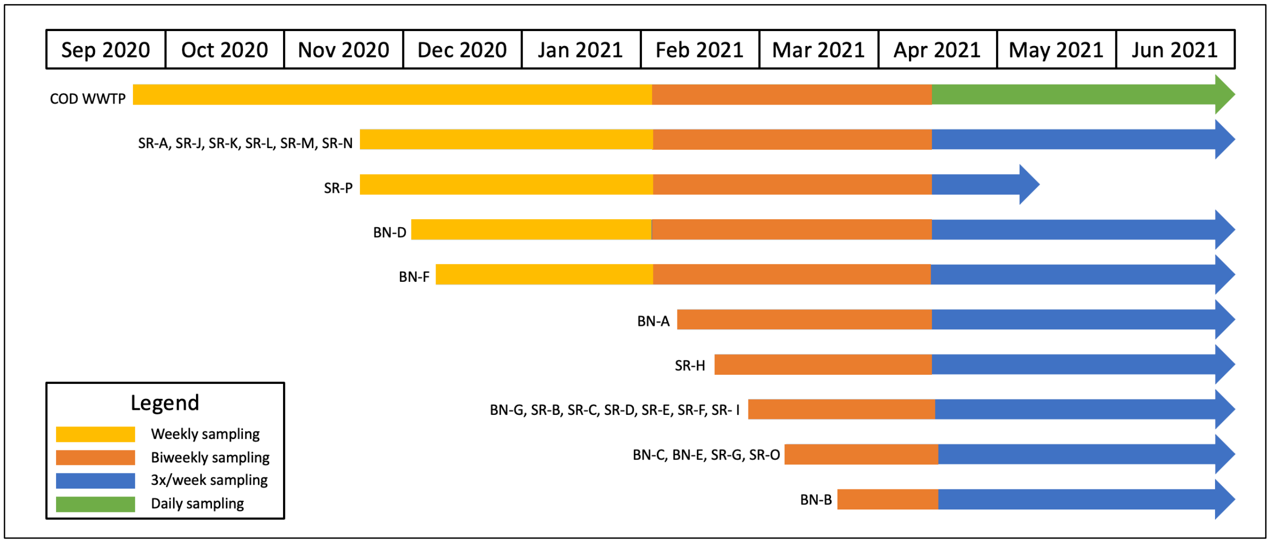


**Figure S1.** Timeline illustrating how zones sampled and sampling frequency evolved over the course of the sampling campaign. Refer to Figure 1 (main text) for locations of zones at the sub-regional (SR) and building/neighborhood (BN) scales.

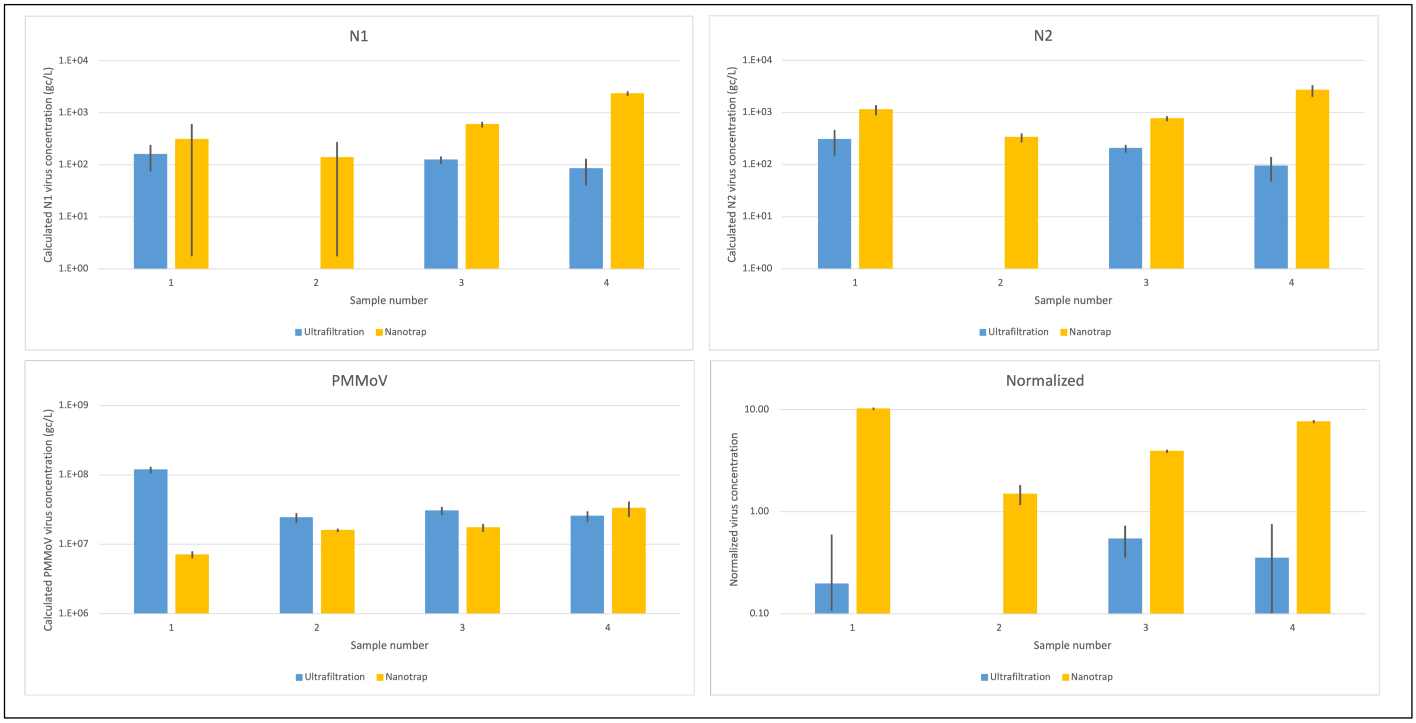


**Figure S2.** Methods comparison results. Bars represent standard error.

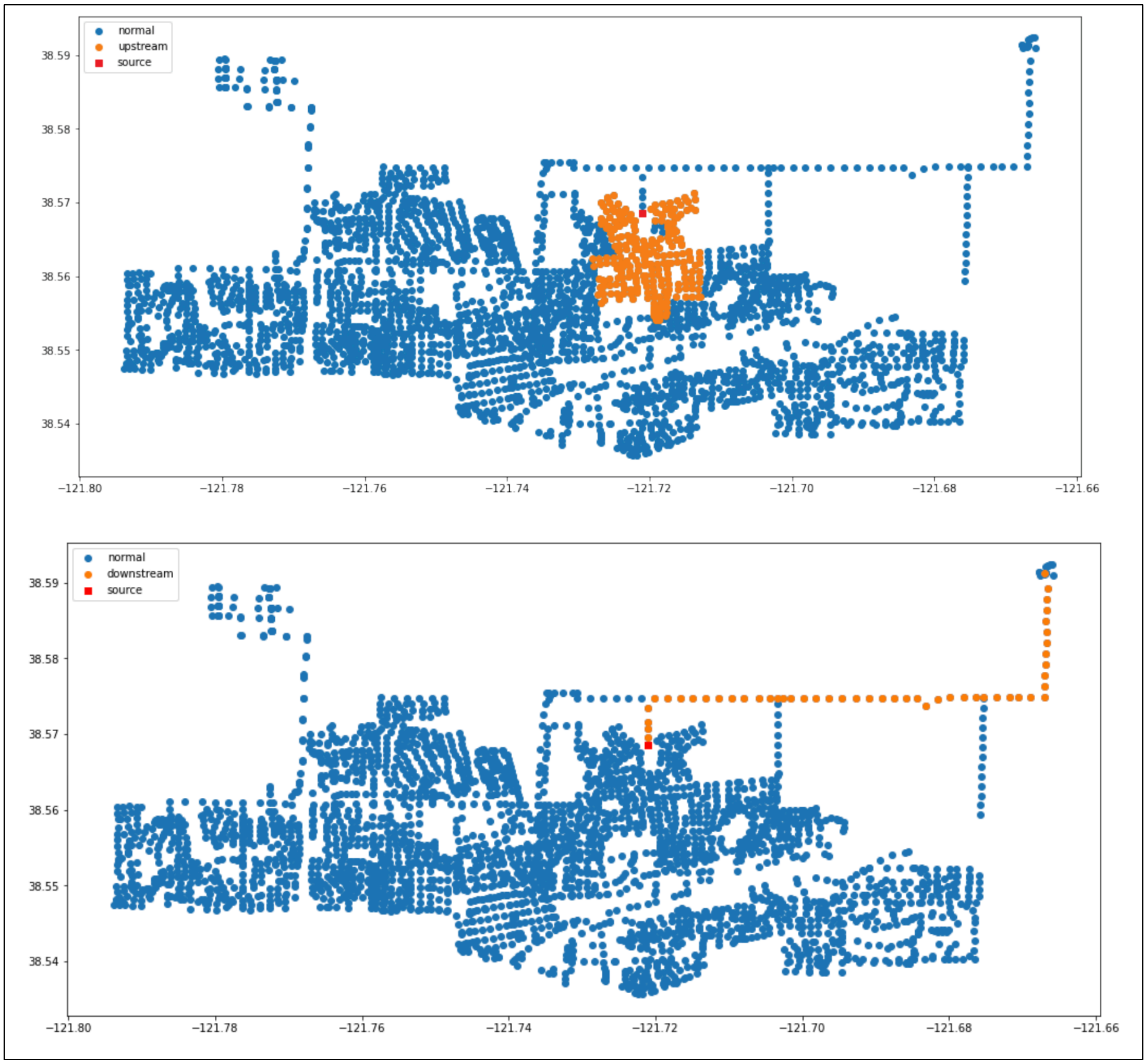


**Figure S3.** Visualization of the connection graph showing all maintenance holes (MHs) in the City of Davis sewershed. At top, orange dots indicate all MHs upstream of a target MH (in red). At bottom, orange dots indicate all MHs downstream of the same target MH.

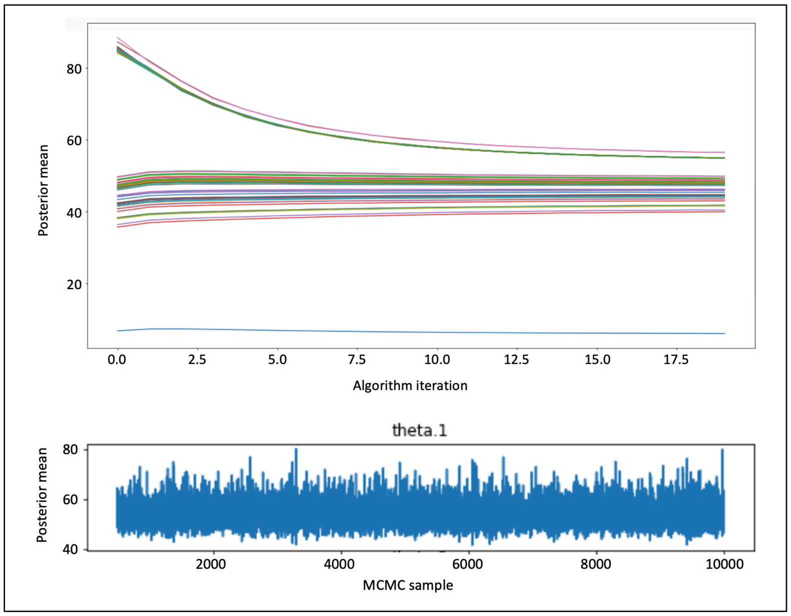


**Figure S4.** Representative quality-check trace plots generated by running the EM-MCMC model on raw qPCR data for Zone SR-L. Top plot illustrates convergence of posterior means over 20 model iterations; colored lines represent posterior means for different sampling dates. Bottom plot illustrates the lack of patterns in MCMC sampling, indicating strong mixing of the Markov chain.

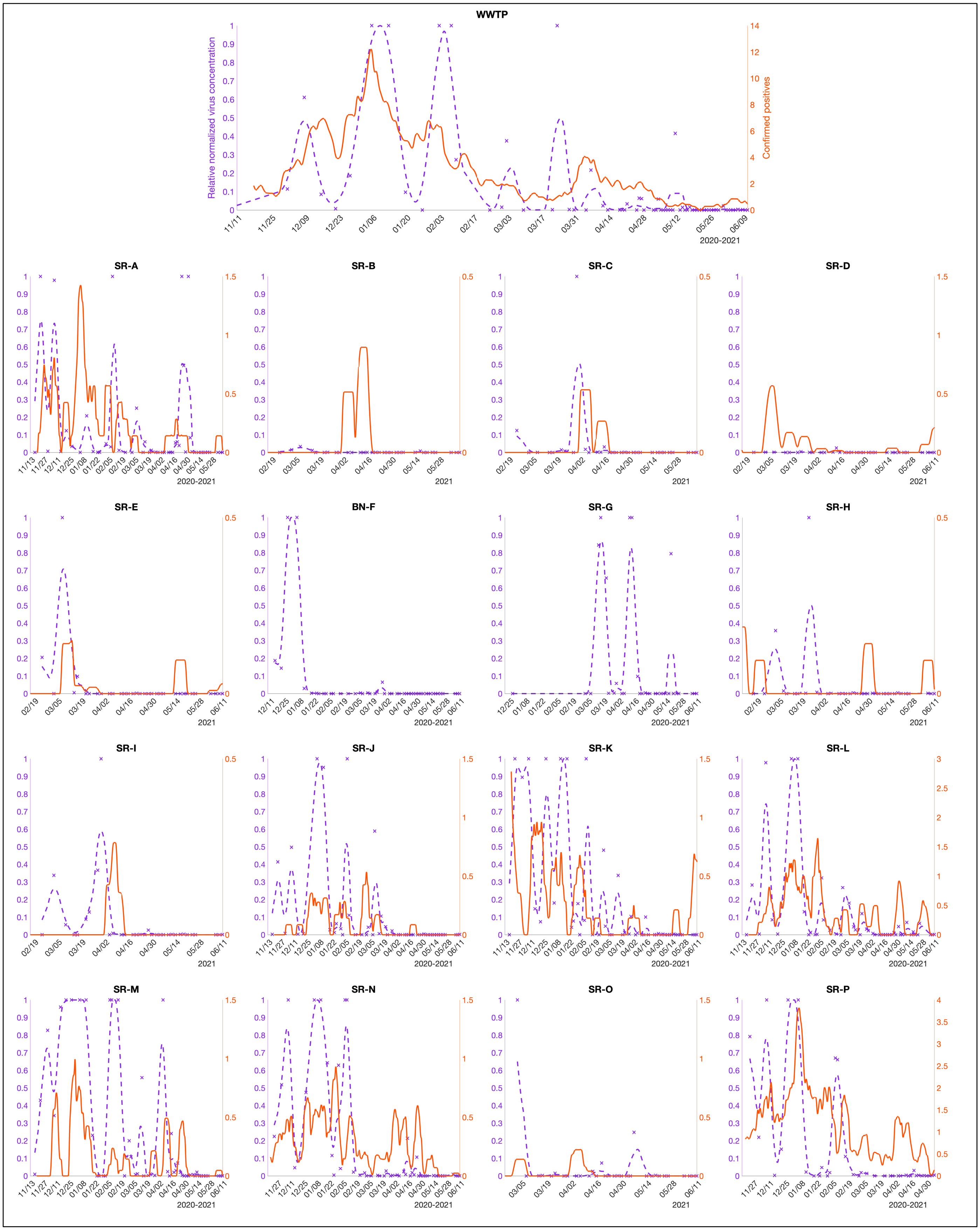
WWTP
Relative normalized virus concentration
Confirmed positives
2020-2021
SR-A
SR-B
SR-C
SR-D
SR-E
BN-F
SR-G
SR-H
SR-I
SR-J
SR-K
SR-L
SR-M
SR-N
SR-O
SR-P
2021

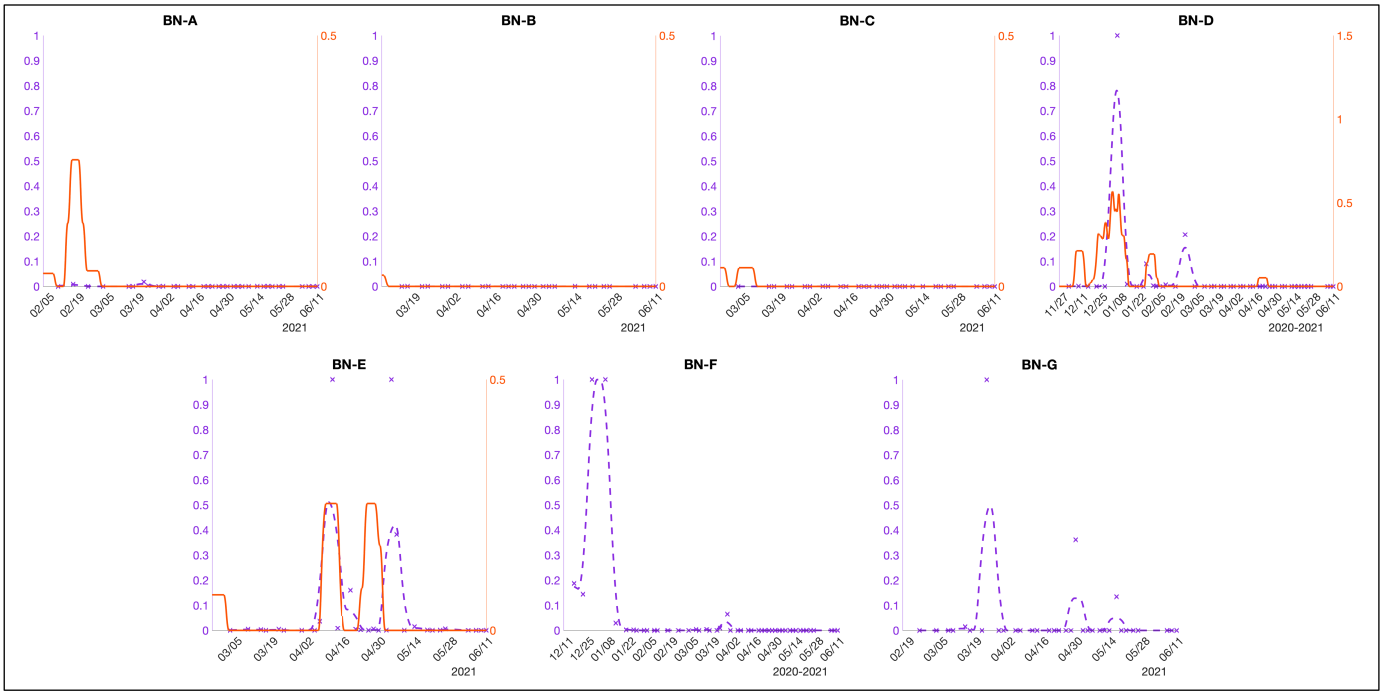


**Figure S5.** Wastewater vs. clinical data in Davis. Xs represent individual sample results; lines represent trends (as centered 7-day moving averages).

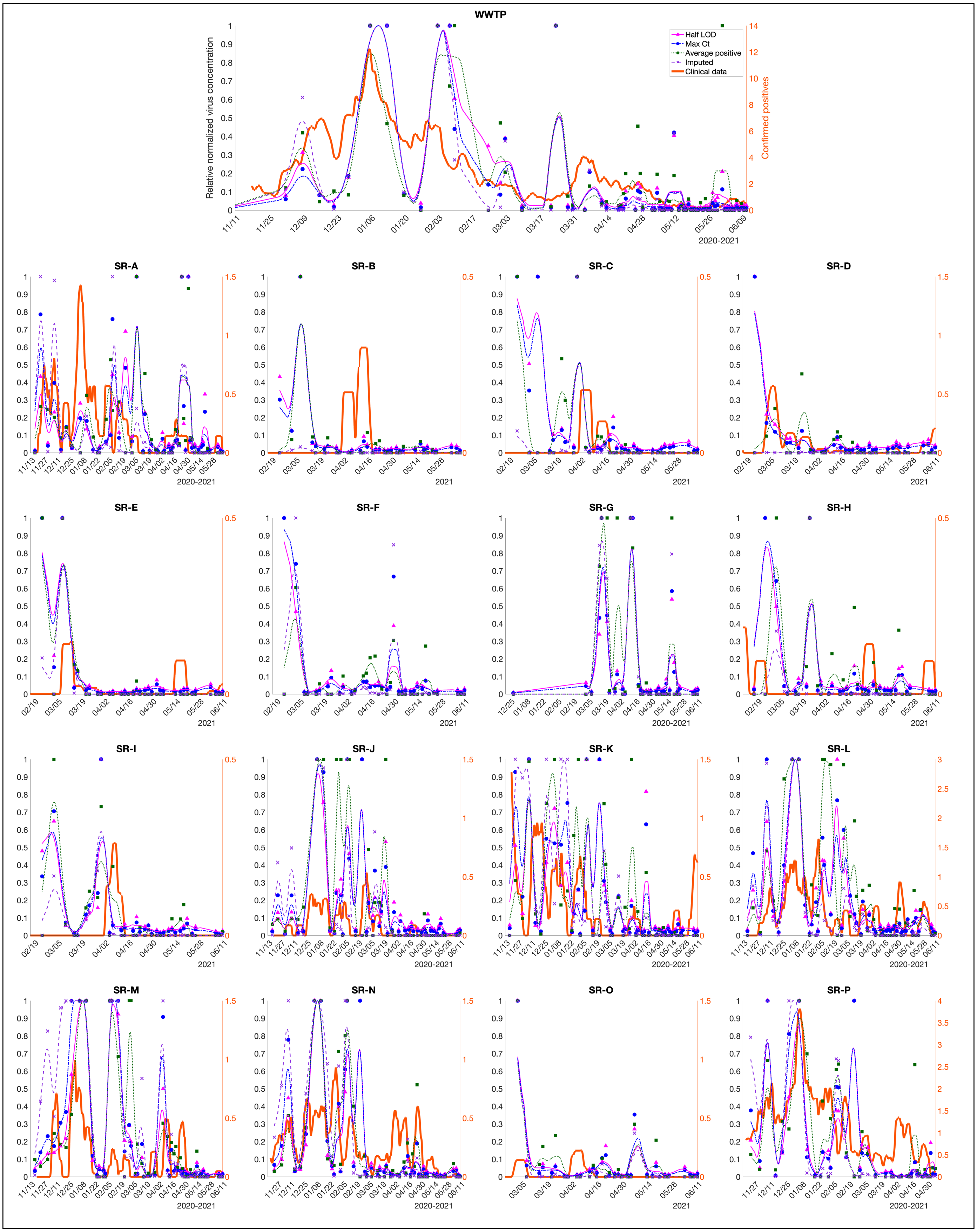
WWTP
Half LOD
Max Ct
Average positive
Imputed
Clinical data
Relative normalized virus concentration
Confirmed positives
2020-2021
SR-A
SR-B
SR-C
SR-D
SR-E
SR-F
SR-G
SR-H
SR-I
SR-J
SR-K
SR-L
SR-M
SR-N
SR-O
SR-P
2021

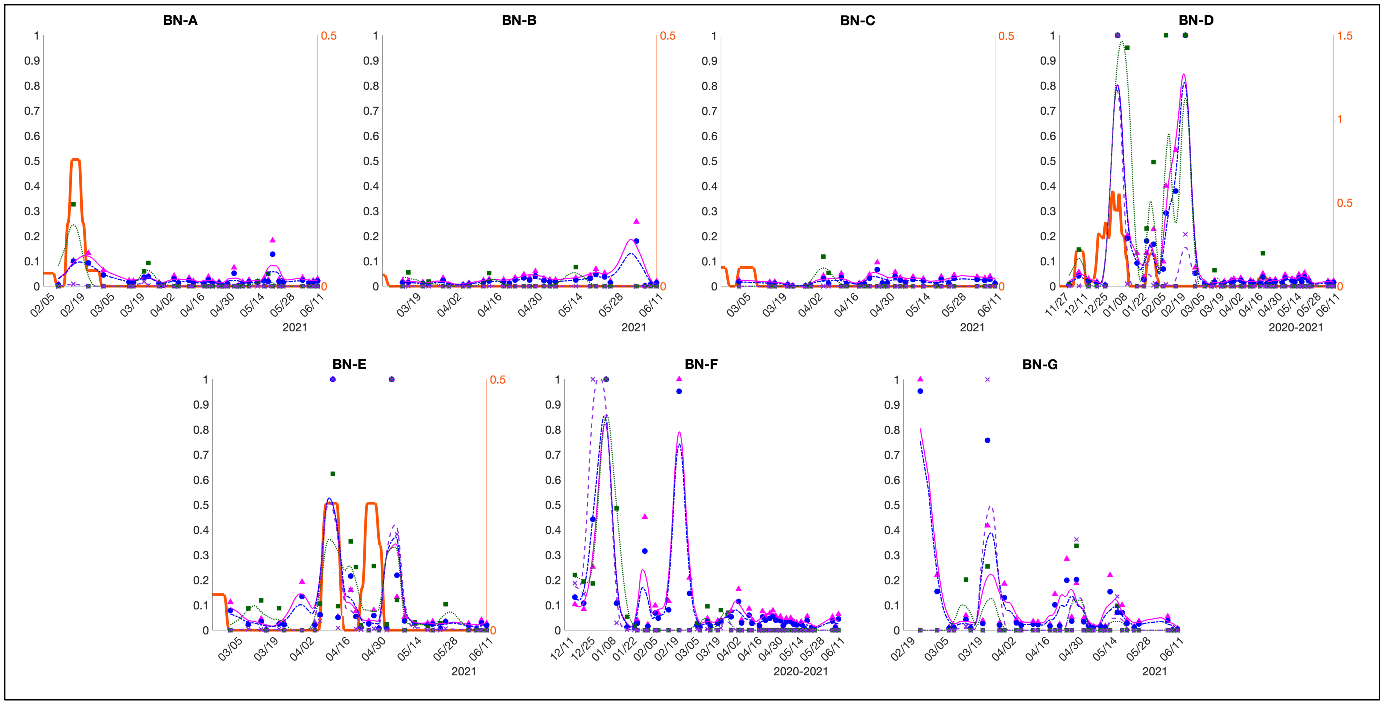


**Figure S6.** Wastewater vs. clinical data in Davis, showing effects of different methods of handling non-detects. Symbols represent individual sample results; lines represent trends (as centered 7-day moving averages).

### Supplementary tables

**Table S1.** Summary of results from methods comparison.

| Sample number | Ultrafiltration | | Magnetic particles | |
|---|---|---|---|---|
| | *Concentration (gc/L)*[*] | *Technical replicates positive (out of 3) per biological replicate* | *Concentration (gc/L)* | *Technical replicates positive (out of 3) per biological replicate* |
| **N1** | | | | |
| 1 | 1.59E+02 (8.40E+01) | 0; 2; 2 | 3.09E+02 (3.07E+02) | 0; 0; 3 |
| 2 | 0 (0) | 0; 0; 0 | 1.39E+02 (1.37E+02) | 0; 0; 3 |
| 3 | 1.25E+02 (1.98E+01) | 1; 2; 2 | 5.99E+02 (7.78E+01) | 3; 3; 3 |
| 4 | 8.54E+01 (4.50E+01) | 0; 1; 2 | 2.36E+03 (2.35 E+02) | 3; 3; 3 |
| **N2** | | | | |
| 1 | 3.05E+02 (1.57E+02) | 2; 2; 3 | 1.14E+03 (2.51E+02) | 3; 3; 3 |
| 2 | 0 (0) | 0; 0; 0 | 3.33E+02 (6.81E+01) | 3; 3; 3 |
| 3 | 2.05E+02 (3.57E+01) | 1; 2; 2 | 7.68E+02 (8.39E+01) | 3; 3; 3 |
| 4 | 9.38E+01 (4.69E+01) | 0; 1; 1 | 2.68E+03 (6.82E+02) | 3; 3; 3 |
| **PMMoV** | | | | |
| 1 | 1.18E+08 (1.29E+07) | 3; 3; 3 | 7.09E+06 (8.32E+05) | 3; 3; 3 |
| 2 | 2.42E+07 (3.99E+06) | 3; 3; 3 | 1.59E+07 (9.30E+05) | 3; 3; 3 |
| 3 | 3.04E+07 (4.28E+06) | 3; 3; 3 | 1.74E+07 (2.26E+06) | 3; 3; 3 |
| 4 | 2.55E+07 (4.50E+06) | 3; 3; 3 | 3.30E+07 (8.28E+06) | 3; 3; 3 |
| **Normalized** | | | | |
| 1 | 0.20 (0.40) | N/A | 10.19 (0.30) | N/A |
| 2 | 0 (0) | N/A | 1.49 (0.33) | N/A |
| 3 | 0.54 (0.19) | N/A | 3.92 (0.15) | N/A |
| 4 | 0.35 (0.40) | N/A | 7.63 (0.29) | N/A |

[*]Upper value indicates average; lower (parenthetical) value indicates standard deviation.

**Table S2.** RT-qPCR primers, probes, and cycling conditions used in this study.

| Target | Primer/probe sequences (5'-3') | | Cycling conditions | Source/Reference |
|---|---|---|---|---|
| SARS-CoV-2; N1 gene | Forward | GACCCCAAAATCAGCGAAAT | 50°C for 30 min; 95°C for 10 min; 45 cycles of 95°C for 15s and 55°C for 45s | U.S. Centers for Disease Control and Prevention (CDC)[4] |
| | Reverse | TCTGGTTACTGCCAGTTGAATCTG | | |
| | Probe | FAM-ACCCCGCATTACGTTTGGTGGACC-BHQ1 | | |
| SARS-CoV-2; N2 gene | Forward | TTACAAACATTGGCCGCAAA | 50°C for 30 min; 95°C for 10 min; 45 cycles of 95°C for 15s and 55°C for 45s | |
| | Reverse | GCGCGACATTCCGAAGAA | | |
| | Probe | FAM-ACAATTTGCCCCCAGCGCTTCAG-BHQ1 | | |
| φ6; P8 protein gene | Forward | TGGCGGCGGTCAAGAG | 50°C for 30 min; 95°C for 10 min; 40 cycles of 95°C for 15s and 60°C for 60s | Gendron et al. (2010)[5] |
| | Reverse | GGATGATTCTCCAGAAGCTGCT | | |
| | Probe | MGB-GTCGCAGGTCTGACACT-BHQ1 | | |
| PMMoV; coat protein gene | Forward | CAGTGGTTTGACCTTAACGTTGA | 50°C for 30 min; 95°C for 10 min; 40 cycles of 95°C for 15s and 60°C for 60s | Zhang et al. (2006)[6] |
| | Reverse | TTGTCGGTTGCAATGCAAGT | | |
| | Probe | MGB-CCTACCGAAGCAAATG-BHQ1 | | |

**Table S3.** Primer/probe mix recipes.

| Target | Recipe | | | |
|---|---|---|---|---|
| | *Reagent* | *Initial concentration* | *Volume added (□L)* | *Final concentration* |
| SARS-CoV-2; N1 and N2 genes | Forward | 100 mM | 13.3 | 500 nM |
| | Reverse | 100 mM | 13.3 | 500 nM |
| | Probe | 100 mM | 3.3 | 125 nM |
| | nuclease-free water | N/A | 170 | N/A |
| φ6 | Forward | 100 mM | 10.7 | 400 nM |
| | Reverse | 100 mM | 10.7 | 400 nM |
| | Probe | 100 mM | 2.1 | 80 nM |
| | nuclease-free water | N/A | 176.5 | N/A |
| PMMoV | Forward | 100 mM | 12.0 | 450 nM |
| | Reverse | 100 mM | 12.0 | 450 nM |
| | Probe | 100 mM | 2.7 | 100 nM |
| | nuclease-free water | N/A | 173.3 | N/A |

[4] CDC. (2021). CDC 2019-Novel Coronavirus (2019-nCoV) Real-Time RT-PCR Diagnostic Panel. Catalog # 2019-nCoVEUA-01.
[5] Gendron, L.; et al. (2010). Evaluation of Filters for the Sampling and Quantification of RNA Phage Aerosols. *Aerosol Science and Technology*, 44: 893–901.
[6] Zhang, T.; et al. (2005). RNA Viral Community in Human Feces: Prevalence of Plant Pathogenic Viruses. *PLoS Biology*, 4(1).

**Table S4.** Master standard curves for each target.

| Target | Standard curve | $R^2$ | Efficiency | Limit of detection (gene copies /reaction)* |
|---|---|---|---|---|
| SARS-CoV-2; N1 gene | y = -3.217x + 38.624 | 0.98 | 104.55% | 0.1 |
| SARS-CoV-2; N2 gene | y = -3.385x + 40.517 | 0.99 | 97.43% | 0.2 |
| φ6 | y = -3.038x + 43.79 | 0.99 | 113.39% | 0.3 |
| PMMoV | y = -3.100x + 40.756 | 0.94 | 110.18% | 153 |

*Reported at a 99% confidence level.

**Table S5.** Number and percent of N1 and N2 non-detects, by sampling scale.

| Sampling scale | N1 | | | N2 | | |
|---|---|---|---|---|---|---|
| | *Total number of non-detects* | *Total technical replicates* | *% non-detects* | *Total number of non-detects* | *Total technical replicates* | *% non-detects* |
| Community | 176 | 231 | 76.2% | 175 | 231 | 75.8% |
| Sub-regional | 1,537 | 1,914 | 80.3% | 1,608 | 1,914 | 84.0% |
| Building/ neighborhood | 686 | 747 | 91.8% | 704 | 747 | 94.2% |

**Table S6.** Average sample Ct, by number of non-detects and average Ct.

| Number of non-detects | Average Ct | |
|---|---|---|
| | *N1* | *N2* |
| 0 | 36.84 | 37.81 |
| 1 | 38.24 | 39.69 |
| 2 | 38.79 | 40.26 |

**Table S5.** Population estimates by sampling zone.*

| Scale | Zone | Population Estimate |
|---|---|---|
| Sub-regional | SR-A | 9,156 |
| | SR-B | 1,076 |
| | SR-C | 1,049 |
| | SR-D | 1,782 |
| | SR-E | 379 |
| | SR-H | 455 |
| | SR-I | 1,030 |
| | SR-J | 1,890 |
| | SR-K | 3,483 |
| | SR-L | 5,048 |
| | SR-M | 1,971 |
| | SR-N | 5,142 |
| | SR-O | 3,949 |
| | SR-P | 24,351 |

| | | |
|---|---|---|
| Building/ neighborhood | BN-A | 496 |
| | BN-B | 147 |
| | BN-C | 624 |
| | BN-D | 1,404 |
| | BN-E | 521 |

*As discussed in the main text, population estimates for Zones SR-F, SR-G, BN-F, and BN-G were unreliable and hence excluded.
